# Associations between outdoor play, screentime and symptoms of anxiety and depression in a non-representative sample of Colombian children: Cross-sectional study

**DOI:** 10.1101/2025.11.03.25339438

**Authors:** Alejandra Tordecilla-Sanders, Gabriela García-Laguna, Andrea-Catalina Trompetero-González, María A. Domínguez-Sanchéz, Diana Marcela Ramos-Caballero, Camila Redondo-Bejarano, Andrés F. Zuluaga-Gómez, Eimy N. Pulecio-Pacheco, Santiago Hernández-Lozano, Eliana Alvarez-Montoya

## Abstract

**Background:** Children’s physical activity (PA) and screentime (ST) levels changed significantly during the pandemic, having negative effects on their mental health, including symptoms of anxiety and depression. No information regarding Colombian children’s PA and ST levels has been reported during the post-pandemic period, or their association with mental health.

**Aim:** Determining associations between outdoor play, screentime and symptoms of anxiety and depression in a non-representative sample of Colombian children.

**Methods:** Data regarding 110 8– to 10-year-old children was subjected to cross-sectional correlational analysis. BMI was determined using weight and height measurements. Mental health was assessed using self-report questionaries for determining Depression and Anxiety symptoms. PA and ST were measured using The Measurement of Physical Activity and Sedentary Behaviour Survey. Bivariate correlational analysis was used and a logistic regression model constructed; statistical significance was determined using <0.05 *p*-value.

**Results:** Overall, 35.4% of children were classified as being overweight or obese. Average indoor playtime was 40.20±38.30 min/day and outdoor 20.78±27.37 min/day. Children spent an average of 125.38±120.39 min/day on screentime. Around 33% of the children had mild and severe anxiety symptoms and 10% depression symptoms. Anxiety symptoms were negatively-correlated with outdoor playtime (Rho=-0.355, *p*-value=0.02) and positively-correlated with depression symptoms (Rho=0.491, *p*-value<0.001). The logistic regression model showed that BMI (OR=0.76, 95% CI: 0.61-0.93) and outdoor playtime (OR=0.97, 95% CI: 0.95-0.99) were significant predictors of reducing the likelihood of moderate-to-severe anxiety symptoms. Depression symptoms were a predictor of the increased likelihood of moderate-to-severe anxiety symptoms (OR=1.41, 95% CI: 1.14-1.88).

**Conclusion:** Outdoor playtime and BMI were associated with target population children’s better mental health. Each additional minute of outdoor playtime per day decreased anxiety symptoms by 3% and each 1kg/m^2^ of BMI by 24%. Depressive symptoms were linked to increased anxiety. PA recommendations based on playtime could promote physical and mental health benefits for children.

## Introduction

The preventive isolation measures adopted for controlling the spread of SARS-CoV-2 had differing effects on physical and mental health associated with changes in lifestyle for the world’s entire population.(1) Some restrictive measures in Colombia involved closing primary and secondary schools, parks, public places and entertainment centres; this meant that educational institutions had to cancel academic classes and sporting activities. Even so, many institutions manged the transition to virtual education.(2)

Many children may thus have had limited access to opportunities for physical activity (PA) during lockdown; engaging in sedentary behaviour, specifically screentime (ST), increased by many hours per day in this population, whilst active playtime decreased.(3) This resulted in various consequences regarding children’s physical and mental health. Evidence shows that isolation had a negative effect on children’s mental health due to closed schools, stress and anxiety at home, along with decreased access to health services.(4–6)

The World Health Organization (WHO) determined that, “One in seven 10-19-year-olds experiences a mental disorder,”(7) stating that, “Depression, anxiety and behavioural disorders are the primary cause of illness and disability among adolescents.”(7) The Colombian Institute of Neurosciences established that 88% of children surveyed showed signs of mental and behavioural health issues.(8) Some studies have described associations between children and adolescents’ sedentary time (specifically regarding ST) and mental health problems related to various risk factors such as lack of proper sleep-time, bad eating habits, dissatisfaction with body weight and cyberbullying, all of which increased during the pandemic.(9,10)

Several strategies have been advanced for promoting healthy habits in children and adolescents for reducing the risk of non-communicable diseases, the main approach being via PA.(11) The WHO has established guidelines on PA and sedentary behaviour patterns which focus on promoting 60 minutes’ minimal time for PA per day, limiting sedentary time (particularly re ST) during recreational periods and taking breaks from sedentary activities every 30 minutes for 6-to 17-year-old children.(12)

Play is universally recognized as one of children’s main activities and is considered essential for their normal growth and development, as it offers physical, emotional, cognitive and social benefits. Furthermore, children come to know their environment and explore the world through play.(13,14) PA has been associated with children and adolescents’ positive mental health effects, including enhanced self-esteem, academic ability, reduced depression and anxiety symptoms and improved cognitive function.(9,10,15–19)

Nutritional Survey of the Nutritional Situation of Colombia (ENSIN)(20) and Physical Activity Report Card(21) surveys have stated that 51% of Colombian children and adolescents meet their PA recommendations, 35.5% achieve PA recommendations and 66.5% spend more than 2 hours on screen activities per day. Furthermore, if a child lives in an urban area, such percentage increases to 71%. No recent reports have been found concerning mental health symptoms and their association with PA, playtime and sedentary time after the pandemic, especially concerning ST in children. This study was thus aimed at determining associations between outdoor play, screentime and symptoms of anxiety and depression in a non-representative sample of Colombian children.

## Materials and methods

### Study design and participants

This study involved a descriptive and correlation analysis as part of a project entitled, *“A comprehensive intervention programme for mitigating the effects of health measures adopted in the city of Bogotá for containing Covid-19 on schoolchildren’s sedentary, emotional and attentional /functional behaviour”.* Such analysis correlated depression and anxiety symptoms with PA (outdoor and indoor playtime) and sedentary time as screentime (computer, videogames and TV) in children from three schools in Bogotá, Colombia after the pandemic, selected by convenience sampling. The study involved 110 8-to 10-year-old children whose parents agreed to them participating. Children suffering cognitive deficiencies, mental, cardiovascular or musculoskeletal disease were excluded. The Universidad del Rosario’s ethics committee approved the research (code DVO0051796-CV1465).

### Measures and procedure

The research team held a meeting with parents and school directors in each school to present the project and obtain their written informed consent; written informed assent was then obtained from each child. The recruitment period for the study was from June 6, 2022 to June 30, 2023. Weight and height measurements were taken at a later date at the three schools, using a stadiometer (SECA 213®, Hamburg, Germany, 1 mm precision, 20-205 cm range) and scales (Tanita Body Fat Monitor for children BF-689®, Tokyo, Japan, 0.1 kg precision, 0-150kg range).

The children’s weight and height were measured according to International Society for the Advancement of Kinanthropometry (ISAK) standards.(22) Children stood on the centre of the scales wearing light clothing and no shoes. Height was measured in the same position, with feet together, the head placed in the Frankfort plane and the stadiometer in contact with the vertex. Body mass index (BMI) was calculated from weight and height data using weight/height (kg/m^2^) formula and classified according to Colombian Ministry of Health and Social Protection age percentile determination(23): <-2 SD underweight, ≥-2 to <-1 SD risk of being underweight, ≥-1 to ≤+1 healthy weight SD, >+1 to ≤+2 SD overweight, >+2 SD obesity.

Depression and anxiety symptoms questionnaires were used in a classroom in optimal conditions (i.e. without sound or visual distractions). The Beck Anxiety Inventory for Children and Adolescents – 2^nd^ version (BAYI-2) was used for assessing anxiety symptoms and the Children’s Depression Inventory – short version (CDI-S) for depression symptoms. The questionnaires were issued and supervised by the researchers’ assistants and the project psychologist.

BAYI-2 was selected for measuring fear and concern associated with anxiety symptoms in 7-to 18-yearold children; it was chosen for its internal consistency (Cronbach’s α 0.62 *p*<0.01) the Spanish version having been validated.(24) This questionnaire consists of 21 questions scored on a Likert-type scale from 0 (never) to 3 (always). Total scores range from 0 to 63, scores being categorised as: minimum (0-7 points), mild (8-15 points), moderate (16-25 points) and severe anxiety symptoms (26-63 points).

The CDI was used for evaluating cognitive, affective and behavioural signs of depression in 7-to 17-year-old children and adolescents. It was selected for its internal consistency (Cronbach α 0.83 *p*<0.01) and has been validated in a Spanish version.(25) The CDI-S assesses such domains through 10 questions scored on a Likert-type scale, indicating 0 for absence, 1 for mild and 2 for high intensity or frequency of depression symptoms. Total score is classified into two categories: absence of symptoms (≤7 points) and presence of depression symptoms (>7 points).

The Measurement of Physical Activity and Sedentary Behaviour Survey was used for measuring sedentary and physical activity time; it was selected for its validity and reproducibility regarding Colombian children (κ-Coefficient for outdoor playtime: 0.31, 95% CI: 0.11-0.48; TV time: 0.52, 95% CI: 0.38-0.64, computer time: 0.70 95% CI: 0.59-0.78, video game time: 0.32 95% CI:0.15-0.47).(26) The questionnaire assesses the time (in minutes) spent on PA (i.e. walking, dancing, biking, swimming, outdoor and indoor playtime) and sedentary activities (sleep time, ST and extracurricular classes) over the past seven days. This questionnaire was digitally completed by parents or caregivers.

The study’s questionnaire items regarding PA included outdoor and indoor playtime on weekdays and at weekends, whilst ST items included time spent on TV, computer and videogames on weekdays and at the weekends. Children were classified according to WHO recommendations for PA as follows: below recommendations (<60 min/day), meeting recommendations (61-120 min/day) and over recommendation (>121 min/day). ST exposure was classified as: meeting recommendations (<120 min/day), over recommendations (120-180 min/day) and well above recommendations (>181 min/day).

### Statistical analysis

Categoric variables were presented as frequencies and percentages whilst continuous variables were reported as means and standard deviations (SD). Descriptive analysis comparing girls and boys involved using the Chi-square test for percentages and either Student’s t-test or the Mann-Whitney U-test, depending on variable normality. Spearman’s rank correlation coefficient was used for assessing PA and ST bivariate correlation with depression and anxiety symptoms. Such analysis was stratified based on weekday and weekend time for PA and ST.

A logistic regression model was built with anxiety symptoms as dependent variable (assuming the presence or absence of moderate to severe symptoms) and depression, BMI, PA and ST as covariables. The model that best fit the data with the least amount of variables was selected, according to Akaike Information Criterion (AIC), using a backward stepwise regression search. R Studio version 4.3.3 (Posit Software, Boston, MA, USA) was used for all statistical analysis; significance level was set at *p*<0.05.

### Missing data

Some ST and PA data was missing due to parents or caregivers choosing not to respond to certain items (Table 1). This meant that correlation and model analysis only concerned close to 70% of the full sample.

**Table 1.** Characteristics, mental health, physical activity and sedentary time by gender.

| Variable | Total<br>(n=110) | Female<br>(n=60) | Male<br>(n=50) | p-value |
| --- | --- | --- | --- | --- |
| General characteristics |  |  |  |  |
| Age, years, mean (SD) | 9.39 (0.85) | 9.33 (0.91) | 9.46 (0.78) | 0.681 |
| Weight, kg, mean (SD) | 34.06 (8.10) | 33.75 (8.78) | 34.42 (7.27) | 0.731 |
| Height, m, mean (SD) | 1.36 (0.08) | 1.36 (0.09) | 1.36 (0.07) | 0.859 |
| BMI, kg/m<sup>2</sup>, mean (SD) | 18.05 (2.78) | 17.88 (2.69) | 18.26 (2.90) | 0.592 |
| BMI classification |  |  |  |  |
| Underweight, n (%) | 1 (0.9) | 1 (1.7) | 0 (0.0) | 0.266 |
| Risk of being underweight, n (%) | 5 (4.5) | 3 (5.0) | 2 (2.4) |  |
| Healthy weight, n (%) | 65 (59.1) | 38 (63.3) | 27 (54.0) |  |
| Being overweight, n (%) | 25 (22.7) | 14 (23.3) | 11 (22.0) |  |
| Being obese, n (%) | 14 (12.7) | 4 (6.7) | 10 (20.0) |  |
| Mental health |  |  |  |  |
| Anxiety, points, mean (SD) | 20.27 (10.25) | 20.22 (9.37) | 20.34 (11.31) | 0.746 |
| Anxiety classification |  |  |  |  |
| Minimum, n (%) | 9 (8.2) | 5 (8.3) | 4 (8.0) | 0.547 |
| Mild, n (%) | 36 (32.7) | 17 (28.3) | 19 (38.0) |  |
| Moderate, n (%) | 29 (26.4) | 19 (31.7) | 10 (20.0) |  |
| Severe, n (%) | 36 (32.7) | 19 (31.7) | 17 (34.0) |  |
| Depression, points, mean (SD) | 3.25 (3.66) | 3.08 (3.33) | 3.46 (4.04) | 0.767 |
| Depression classification |  |  |  |  |
| Absence of depression, n (%) | 99 (90.0) | 55 (91.7) | 44 (88.0) | 0.537 |
| Presence of depression, n (%) | 11 (10.0) | 5 (8.3) | 6 (12.0) |  |
| Physical activity |  |  |  |  |
| Play indoors, min/day, mean (SD) | 40.20 (38.30) | 38.65 (40.45) | 42.12 (35.98) | 0.330 |
| Play indoors weekdays, min/d, mean (SD) | 37.77 (36.28) | 34.95 (41.02) | 41.27 (29.61) | 0.077 |
| Play indoors weekends, min/d, mean (SD) | 46.28 (64.67) | 47.92 (64.66) | 44.24 (65.63) | 0.733 |
| Play outdoors, min/day, mean (SD) | 20.78 (27.37) | 21.46 (24.61) | 19.94 (30.82) | 0.668 |
| Play outdoors weekdays, min/d, mean (SD) | 14.89 (26.25) | 15.53 (23.34) | 14.10 (29.83) | 0.849 |
| Play outdoors weekends, min/d, mean (SD) | 35.51 (51.22) | 36.28 (53.91) | 34.56 (48.47) | 0.872 |
| Physical activity classification |  |  |  |  |
| Below recommendations, n (%) | 9 (8.2) | 6 (10.0) | 3 (6.0) | 0.719 |
| Meeting recommendations, n (%) | 20 (18.2) | 9 (15.0) | 11 (22.0) |  |
| Over recommendations, n (%) | 46 (41.8) | 26 (43.3) | 20 (40.0) |  |
| No data, n (%) | 35 (31.8) | 19 (31.7) | 16 (32.0) |  |
| Sedentary time |  |  |  |  |
| Screen time, min/d, mean (SD) | 125.38 (120.39) | 133.33 (121.81) | 115.84 (119.19) | 0.496 |
| Screentime weekday, min/d mean (SD) | 85.07 (121.69) | 84.0 (123.43) | 86.36 (120.80) | 0.395 |
| Screentime weekend, min/d mean (SD) | 105.22 (114.75) | 108.66 (114.26) | 101.10 (116.35) | 0.935 |
| Screentime classification |  |  |  |  |
| Meeting recommendations, n (%) | 53 (48.2) | 28 (46.7) | 25 (50.0) | 0.410 |
| Over recommendations, n (%) | 9 (8.2) | 7 (11.7) | 2 (4.0) |  |
| Well above recommendations, n (%) | 10 (9.1) | 4 (6.7) | 6 (12.0) |  |
| No data, n (%) | 38 (34.5) | 21 (35.0) | 17 (34.0) |  |
| Abbreviations: %= percentage by group; min/d= minutes per day; n= number of children; SD= standard deviation |  |  |  |  |

## Results

Girls accounted for 54.5% of the children; mean age was 9.39±0.85 years and BMI 18.05±2.78 kg/m^2^. Regarding weight, 59% of the children were catalogued as having healthy weight and 35.4% as being overweight and obese; the data highlighted a greater prevalence for girls being overweight (23.3%) than boys (22%) and higher prevalence for boys being obese (20.0%) than girls (6.7%). Average indoor playtime was 40.20±38.30 min/day and outdoor PA 20.78±27.37 min/day, more time being spent on weekends for both modalities (46.28±64.67 min/day indoor playtime *cf* 35.51±51.22 min/day outdoor playtime).

Children spent 125.38±120.39 min/day on ST, being higher on the weekends than weekdays (105.22±114.75 min/day *cf* 85.07±121.69 min/day). No significant differences were found regarding gender, playtime (outdoor and indoor) and screentime regarding anxiety and depression symptoms (Table 1).

Anxiety symptoms mean score was 20.27±10.25, about 33% of the children having mild and/or severe symptoms. Boys had greater prevalence for anxiety symptoms (34%) than girls (31.1%); 90% did not exhibit depression symptoms. A similar trend was found regarding greater depression symptoms in boys (12%) than girls (8.3%) (Table 1).

Correlating anxiety symptoms with PA (indoor and outdoor playtime) and ST (TV, video game and computer time) per day, our study found that anxiety symptoms had a weak-to-moderate inverse correlation with outdoor playtime (Rho=-0.355; *p*=0.02), as shown in figure 1. A moderate correlation was found between anxiety and depression symptoms (Rho=0.491, *p*-value<0.001). No correlation was found between screentime and anxiety or depression symptoms (Fig 1).

**Fig 1.**
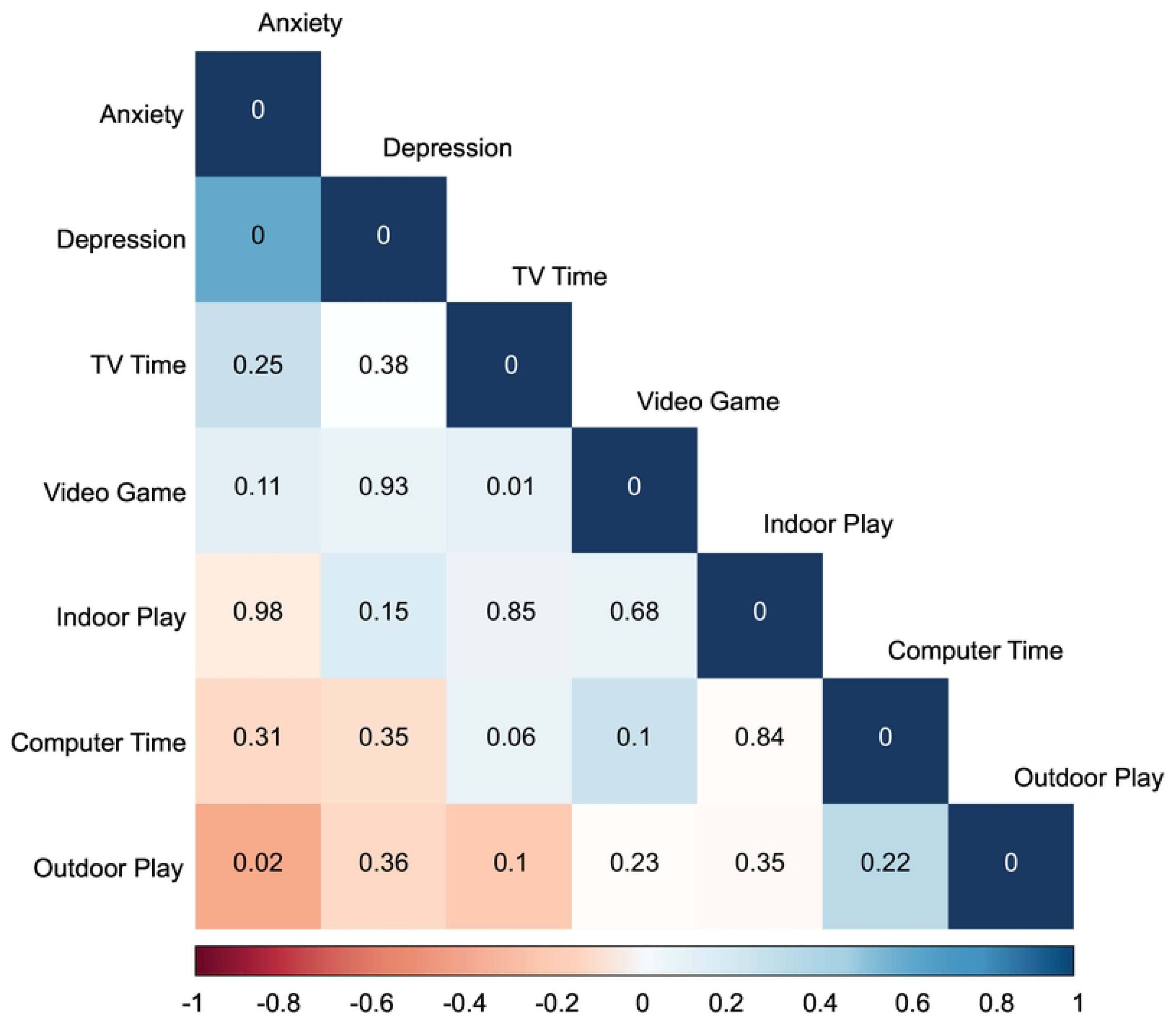
General correlations chart. Anxiety and Depression corresponds to the anxiety and depression symptom soore. lndoor play, Outdoor play, TV Time, Video game and Computer time oorresponds to the average time spent on these activities per day. Values presentend on the chart correspond to p-value of bivariate oorrelations.

It was found that anxiety (Rho=-0.303; p=0.032) and depression symptoms (Rho=0.257; p=0.019) had a week-to-moderate inverse relationship with outdoor playtime on weekdays, when analysing relationships between anxious and depressive symptoms concerning PA and ST on weekdays and weekends. Depression symptoms had a very weak direct correlation with indoor playtime on weekdays (Rho=0.178, p-value=0.028). No statistically significant correlations were found for weekdays or weekends regarding ST. (Fig 2 and 3).

**Fig 2.**
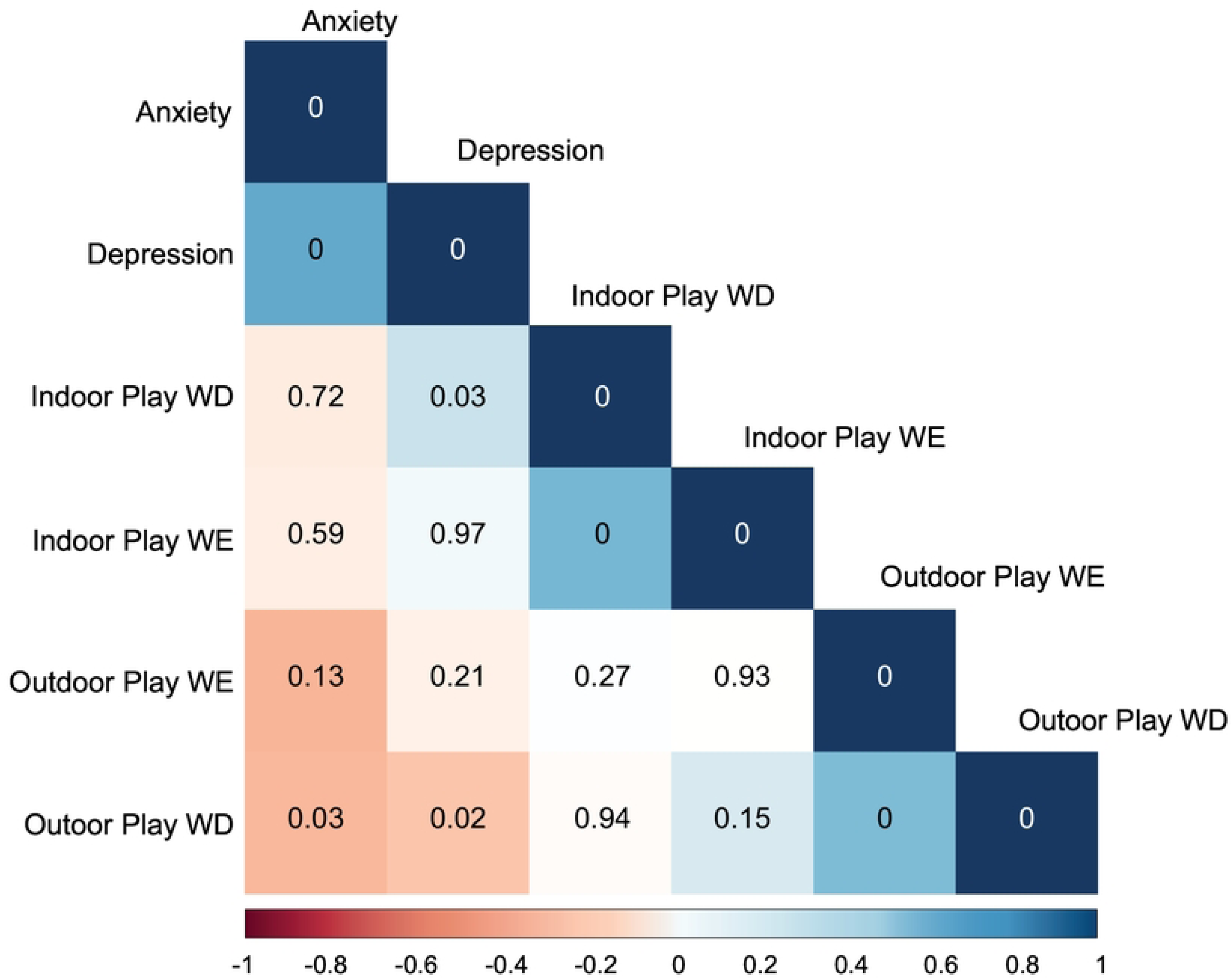
Correlations chart for Mental Health and Physical Activity. Note. Anxiety and Depression corresponds to the anxiety and depression symptom score. Indoor play, Outdoor play time corresponds to the average time spent on these activities per day. Values presentend on the Chart correspond to p-value of bivariate correlations. WD: Weekday, WE: Weekend.

**Fig 3.**
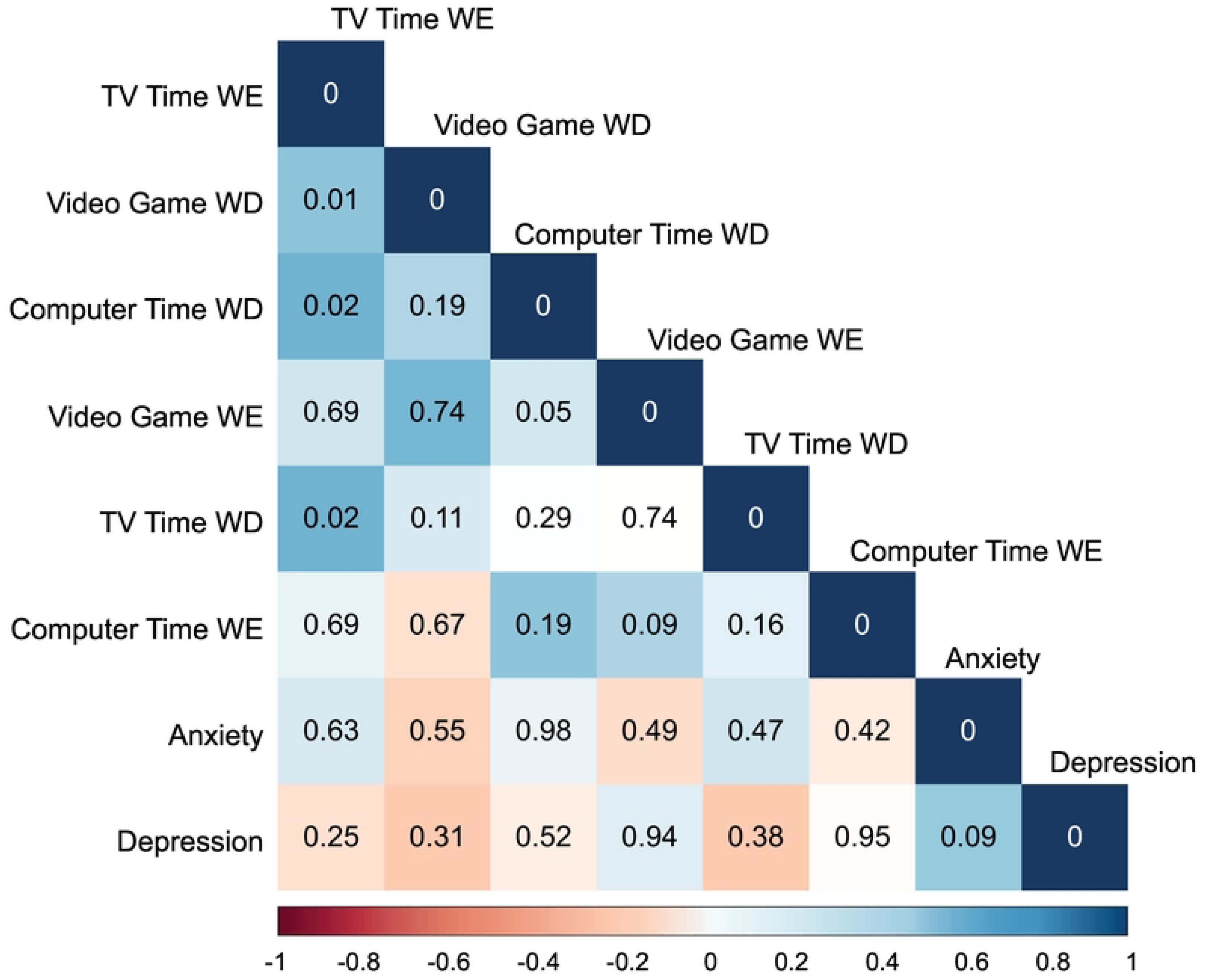
Correlations chart for Mental Health and Screen Time. Note. Anxiety and Depression corresponds to the anxiety and depression symptom score. TV, Video game and Computer time corresponds to the average time spent on these activities per day. Values presentend on the chart correspond to p-value of bivariate correlations. WD: Weekday, WE: Weekend.

Regarding bivariate correlations results, logistic regression used the presence of moderate-to-severe (1) or minimal-to-mild (0) anxiety symptoms as dependent variable. It was found that BMI (OR=0.76 95%CI: 0.61-0.93) and outdoor playtime (OR=0.97 95%CI: 0.95-0.99) were significant predictors for reducing the likelihood of having moderate-to-severe anxiety symptoms. This meant that the odds of presenting moderate-to-severe anxiety symptoms were 24% for every increase in 1kg/m^2^ re BMI and 3% lower for each additional minute per day of outdoor playtime.

Conversely, depression symptoms were a predictor of an increased likelihood of experiencing moderate-to-severe anxiety symptoms (OR=1.41, 95% CI: 1.14-1.88), indicating that each additional point in depression symptoms was associated with a 41% higher likelihood of experiencing moderate-to-severe anxiety symptoms (Fig 4). No variables regarding ST had a relationship with anxiety symptoms. These results were taken into account since the model explained 42.2% (Nagelkerke R^2^=0.422) of variance regarding anxiety symptoms and the model’s good predictive capability (AUC=0.83, sensitivity=0.872, specificity=0.519) (Fig 5).

**Fig 4.**
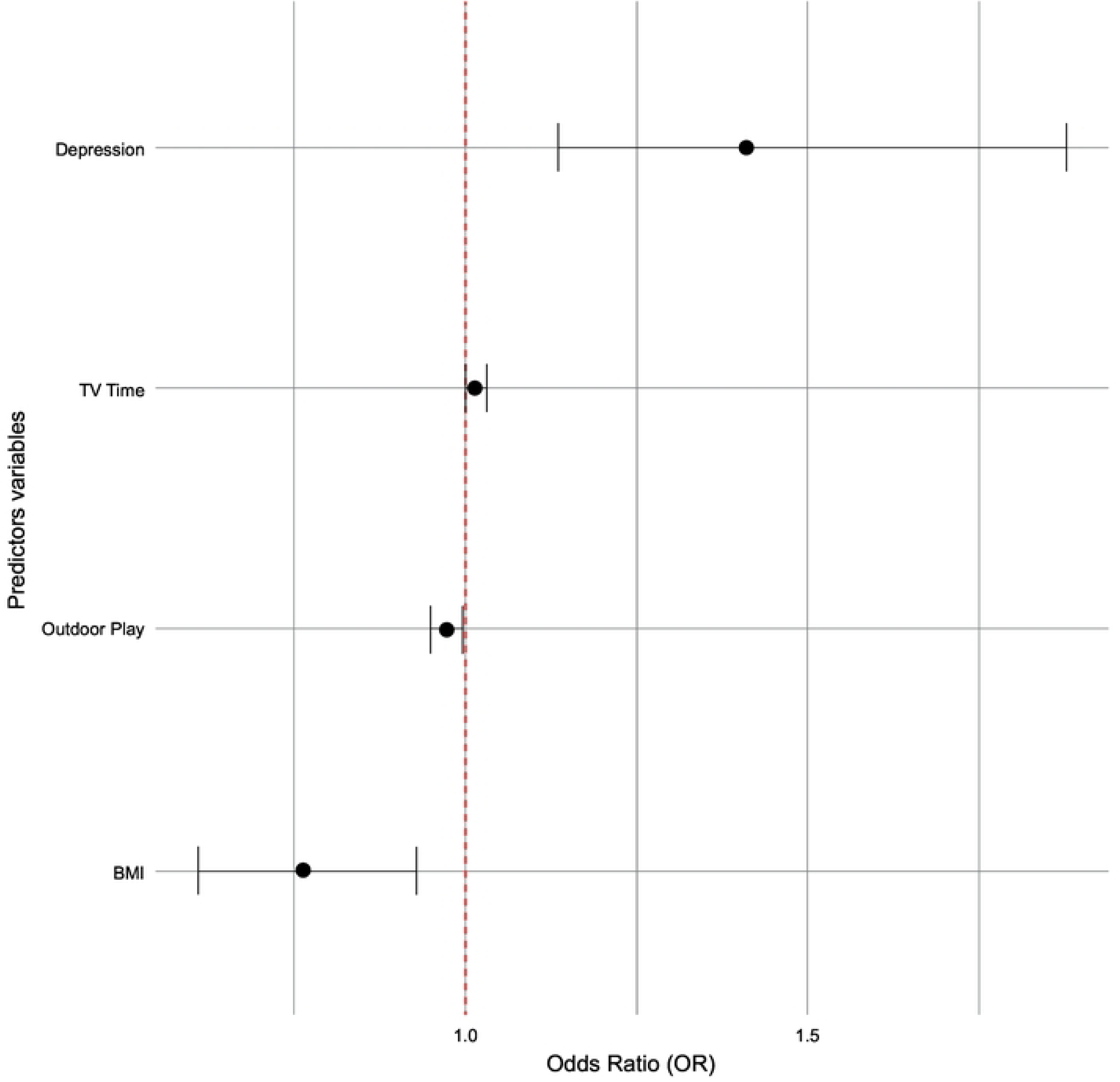
Odds Ratios for the logistic regression model for Anxiety Symptoms. Note. Depression is the score for symptoms, TV time and outdoor playtime is the soore for the average time spent on these activities per day, body msss index (BMI). Points represent the OR and Error bars show the 95% CI for the OR.

**Fig 5.**
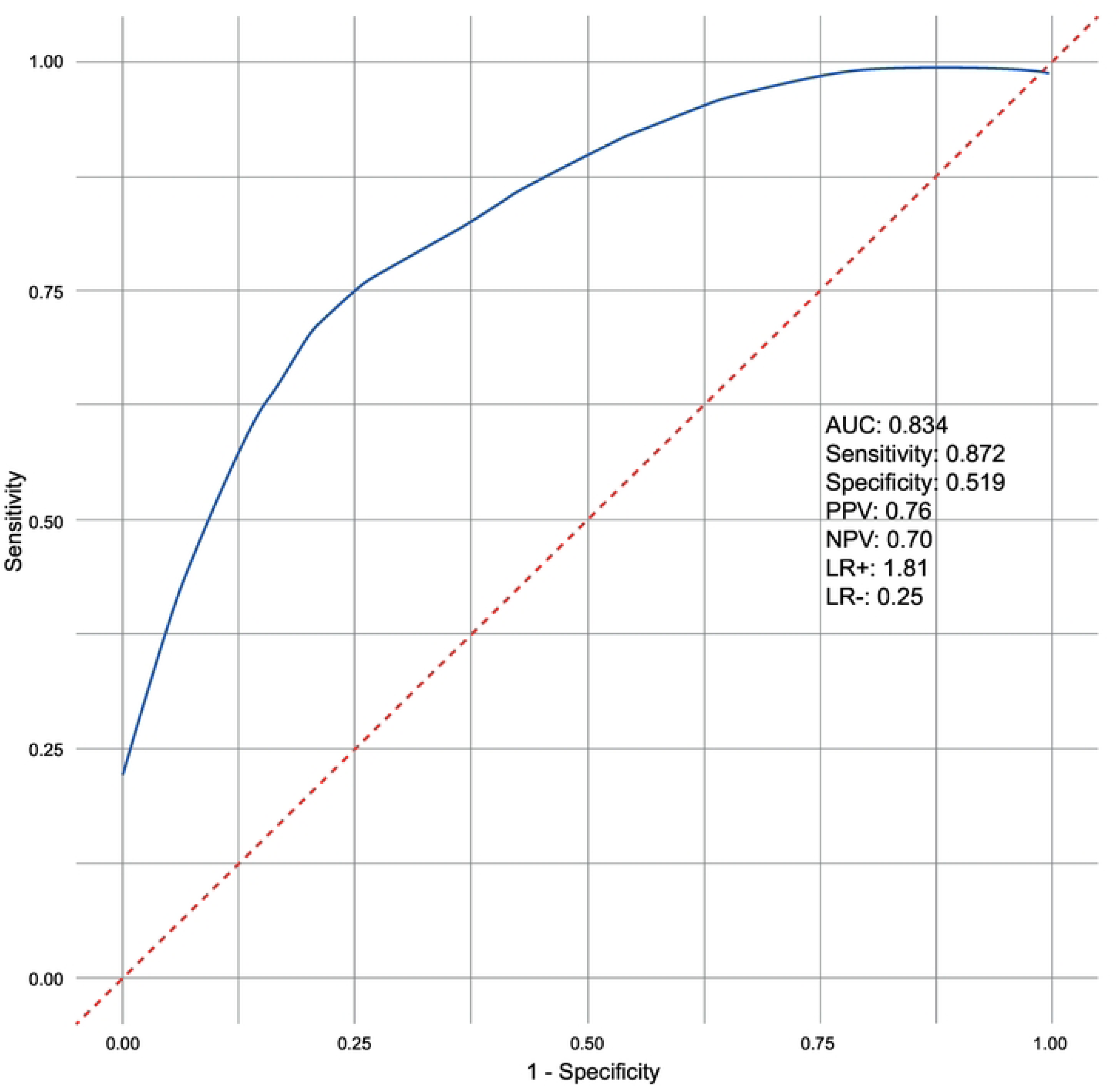
ROC Curve for the Logistic regression model for Anxiety Symptoms. Nota: Note. Area under the curve(AUC), positive predictive values (PPV), negative predictive values (NPV), positive likelihood ratio(LR+), negative likelihood ratio (lR−).

## Discussion

Our findings revealed a high prevalence for children having healthy weight, there being a higher prevalence of obesity in boys than girls; such results were similar to those reported in Nutritional Survey of the Nutritional Situation of Colombia (ENSIN 2015) in which 25.3% of boys and 23.5% of girls were overweight. (20) The study also revealed that 33% of the boys had mild and severe anxiety symptoms. Such findings were similar to those reported by Ghandour et al. (27) stating that 45.2% of children in their study had mild, 47.2% moderate and only 7.6% severe anxiety conditions. Luque et al. (28) reported 15.7% depression symptoms in Colombian children from Barranquilla, agreeing with our finding of 10% using the CDI questionnaire.

Our study found that 60% of the target children achieved PA recommendations regarding outdoor and indoor playtime and that 48.2% spent less than 2 hours per day on ST. The Colombian ENSIN 2015 survey reported that only 31.1% of children spent more than 60 minutes per day in active play and 67% of children spent more than 2 hours per day on ST during their leisure time.(20) Whiting S et al. (29) reported similar results, stating that 4 in 5 European children spent at least 1 hour per day in active play whilst (conversely regarding our findings) 60.2% of children engaged in ST for <2h/day. Our study found that children spent more time on indoor playtime, such results being like those reported by Dodd et al. (30), determining that British children spent more time playing at home during 2020 and most adventurous play happened in green spaces and indoor playcentres.

Our study found that higher anxiety and depression symptoms were correlated with lower time spent in outdoor play, thereby highlighting outdoor playtime’s protective factor regarding anxiety symptoms (OR= 0.97, 95% CI:0.95-0.99). These results were similar to those reported by McMahon et al. (31) who found that adolescents participating in sports was associated with significantly lower levels of anxiety and depressive symptoms for girls and boys (*p*<0.0005).

Our results did not enable determining causality; however, in justifying indoor playtime’s direct weak relationship with depressive symptoms, it was hypothesised that a lack of intrinsic motivation during home time was the possible cause of such symptomatology. Meyer et al. (32) determined that physical activity’s beneficial effect only occurs if it is coupled with intrinsic motivation and activity levels are high. Yanjie et al. (33) reported the direct moderate relationship between anxiety and depression symptoms and their predictor role regarding the increased likelihood of experiencing symptoms (OR= 1.41, 95%CI: 1.14-1.88). Their findings reported an independent association of depressive and anxiety symptoms in primary school children (OR= 4.935, 95% CI: 4.442-5.485).

BMI in the model is a protector factor regarding anxiety symptoms in children (OR= 0.76, 95%CI: 0.61-0.93); nevertheless, evidence has shown a U-shaped association between anxiety and BMI in a white population;(34) being underweight and obese was thus more likely to be associated with anxiety compared to normal weight individuals’ anxiety. Furthermore, other studies have stated and explained that other factors can be strongly associated with anxiety, such as low self-esteem, bodily, environmental, social context and cultural dissatisfaction. (35,36)

No correlation was found between ST and anxiety or depression symptoms; it was not a factor explaining the symptomatology, thereby coinciding with the results of the systematic review by Tang et al. (37), who reported that increased screen time is unlikely to explain the substantial increase in the prevalence of depressive symptoms among young people. On the other hand, our results differ from those reported by Boers et al. (38), who attributes the increase in anxiety and depression rates among adolescents to excessive time spent in front of digital screens. The use of social media, television, and computers is associated with high levels of anxiety. The significant effect within the individual on social media use indicated that a one-hour increase in the average time spent using social media in a given year was associated with an increase in anxiety symptoms (β = 0.21, Std. Er: 0.03, 95% CI: 0.14-0.28). Also, but more modest within-person relationships were revealed for television viewing and computer use on anxiety (β = 0.15, Std. Er: 0.03, 95% CI: 0.08-0.22). In the study of Maras et al. (39), the duration of screen time was associated with severity of depression (β = 0.23, *p* <0.001) and anxiety (β = 0.07, *p* < 0.01). Video game playing (β = 0.13, *p* < 0.001) and computer use (β = 0.17, *p* < 0.001) but not tv viewing were associated with more severe depressive symptoms. Although these findings differ from those found in the present study, it is suggested that teachers, parents, and students be educated on the use of digital devices to prevent impacts on the mental health of children and adolescents and evaluate the effect of these educational strategies on the population. (38)

On the other hand, one of this study’s limitations was the use of a self-report questionnaire to be answered by parents or caregivers for measuring PA and ST; this may have led to varying interpretations by different cultures and age groups. The convenience sampling, small sample size and large amount of missing data regarding PA and ST variables probably led to bias. The data’s cross-sectional nature impeded determining causal relationships between PA, ST and mental health. Further research should explore the longitudinal impact of PA and ST on Colombian children

## Conclusions

Children’s playtime-related PA was associated with better mental health. Each additional minute of outdoor playtime per day decreased children’s anxiety symptoms by 3%; such finding highlights the importance of increasing children’s PA levels and maintaining BMI within normal ranges may help prevent anxiety symptoms. ST was not found to be associated with mental health in target population children. PA recommendations for children should be met by ensuring that accessible play activities are provided. More intervention studies are recommended for enabling cause-effect information concerning the effect of PA on children’s mental health to be obtained.

## Data Availability

The information will only be available after acceptance

## Acknowledgments

We would like to thank the educational institutions which allowed us access to the children, parents or guardians and teachers involved in this study, as well as the project’s statistical advisor and team of evaluators, including psychologists.

The project entitled, “A comprehensive intervention programme for mitigating the effects of health measures adopted in the city of Bogotá for containing Covid-19 on schoolchildren’s sedentary, emotional and attentional/functional behaviour”, had funding from a 2020 initiative involving the Universidad del Rosario and Universidad Nacional de Colombia focused on mitigating COVID-19 effects in Colombia.

## Authors’ contributions

ATS: conceptualisation, methodology, research, project administration, data analysis, manuscript writing (writing, reviewing and editing); GGL, ACTG, MADS, DMRC, SHL: research, manuscript writing (reviewing and editing); AFZG, CRB, ENPP, EAM: data collection, data analysis, manuscript writing (writing, reviewing and editing).

## Competing interests

The authors declare that they have no competing interests.

## Notes

### Competing Interest Statement

The authors have declared no competing interest.

### Funding Statement

Yes

### Author Declarations

The Universidad del Rosario’s ethics committee approved the research (code DVO0051796-CV1465). Written informed consent was obtained from the parents, and informed assent from the children.

